# The spatial spread of HIV in Malawi: An individual-based mathematical model

**DOI:** 10.1101/2020.12.23.20248757

**Authors:** Janne Estill, Wingston Ng’ambi, Liudmila Rozanova, Olivia Keiser

## Abstract

The prevalence of HIV varies greatly between and within countries. We therefore developed a flexible individual-based mathematical model for HIV transmission, that comprises a spatial representation and individual-level determinants. We tested this model by calibrating it to the HIV epidemic in Malawi and exploring whether the heterogeneity in HIV prevalence could be caused without accounting for heterogeneity in behaviour. We ran the model for Malawi between years 1975-2030 with five alternative realizations of the geographical structure and mobility: (I) no geographical structure; 28 administrative districts including (II) only permanent relocations between districts, (III) permanent relocations and between-district casual sexual relationships, or (IV) permanent relocations between districts and to/from abroad and between-district casual sex; and (V) a grid of 10×10km^2^ cells, with permanent relocations and between-cell casual relationships. We assumed HIV was present in 1975 in the districts with >10% prevalence in 2010. We calibrated the models to national and district-level prevalence estimates.

Reaching the national prevalence required all adults to have at least 20 casual sex acts/year until 1990. Models II, III and V reproduced the geographical heterogeneity in prevalence to some extent if between-district relationships were either excluded (Model II) or restricted to minimum (Models III, V). Long-distance casual partnership mixing (Models III-V) mitigated the differences in prevalence substantially; with international migration the differences disappeared completely (Model IV). National prevalence was projected to decrease to 4-5% by 2030. Our model sustained the major differences in HIV prevalence across Malawi, if casual relationships between districts were kept at sufficiently low level. An earlier introduction of HIV into the Southern part of Malawi may thus be one of the explanations to the present heterogeneity in HIV prevalence.

**Author summary:** The prevalence of HIV varies greatly across the settings, both globally and within countries. The ability of the commonly used compartmental models to account for the geographical structure and individual-level determinants that cause this heterogeneity is limited. In this project, we developed an individual-based simulation framework for modelling HIV transmission in a real setting. We built the model to take into account an unlimited number of individual-level characteristics, and a geographical representation of the setting that can be defined using an arbitrary resolution and distance matrices. We demonstrate the use of this model by simulating the HIV epidemic of Malawi 1975-2030 and exploring whether the observed heterogeneity could be preserved without taking into account any spatial heterogeneity in sexual behaviour. A relatively simple version of the model reproduced the broad-scale differences in HIV prevalence, but the detailed differences will need further investigation.

## Introduction

HIV is one of the most serious global health emergencies that occurred during the past few decades. The severity of the HIV epidemic varies greatly across the world. Currently, adult HIV prevalence in the southernmost countries of Africa ranges between 9% and 27%, whereas outside of Africa there is no country with a prevalence above 2%.[1] Differences exist also within countries.[2] In 2019, Malawi had a national HIV prevalence of 8.9% with the district-level prevalence ranging between 4% and 22% and the epidemic being more severe in the southern part of the country. [3]

The reasons behind the variability remain disputed. The risk of HIV depends on a complex network of biomedical, socioeconomic, cultural and behavioural factors and geographical structure.[4] People living in densely populated or well-connected areas may have a broader contact network, and thus the likelihood to have at least one partner who is infected is higher. International connectivity may also play a role. It is however unclear to what extent the varying burden of HIV is a result from the times and locations where HIV was first introduced into the country and the connections between the different geographical locations.

Mathematical models are an essential tool in the evaluation of dynamics of infectious disease epidemics. We developed an individual-based mathematical model including a geographical structure to simulate the course of the Malawian HIV epidemic between 1975 and 2030. In the present study, we describe the technical details of the model, test if the observed variability in HIV prevalence can be reproduced by differences in the early stages of the epidemic, and make future projections for district-level HIV prevalence.

## Methods

### Model description

Our individual-based model consists of two interacting modules (Fig. 1; <u>Text S1, Table S1</u>; the full model code is available on https://gitlab.com/igh-idmm-public/agent-based-hiv-transmission-model). The *transmission module* represents the population of the desired setting (Malawi). At the beginning of the simulation, an initial population is generated. For each individual we sample his/her age, sex, location of residence, a set of behavioural characteristics and biomedical factors, and the HIV status. Each child under age 15 is assigned a woman aged >15 years as mother, and men and women are paired to form regular partnerships. The population is updated in steps of one year, when the following events are sampled from pre-defined probabilities: an individual or family may move to another location; an individual’s socio-behavioural or biomedical characteristics may change; a couple may separate and single individuals may form partnerships; uninfected individuals may acquire HIV; an individual may die; and infants may be born to women of childbearing age. The number of locations in the model can be chosen arbitrarily, and separate distance matrices are defined for permanent relocation and for selecting casual partners.

**Figure 1.**
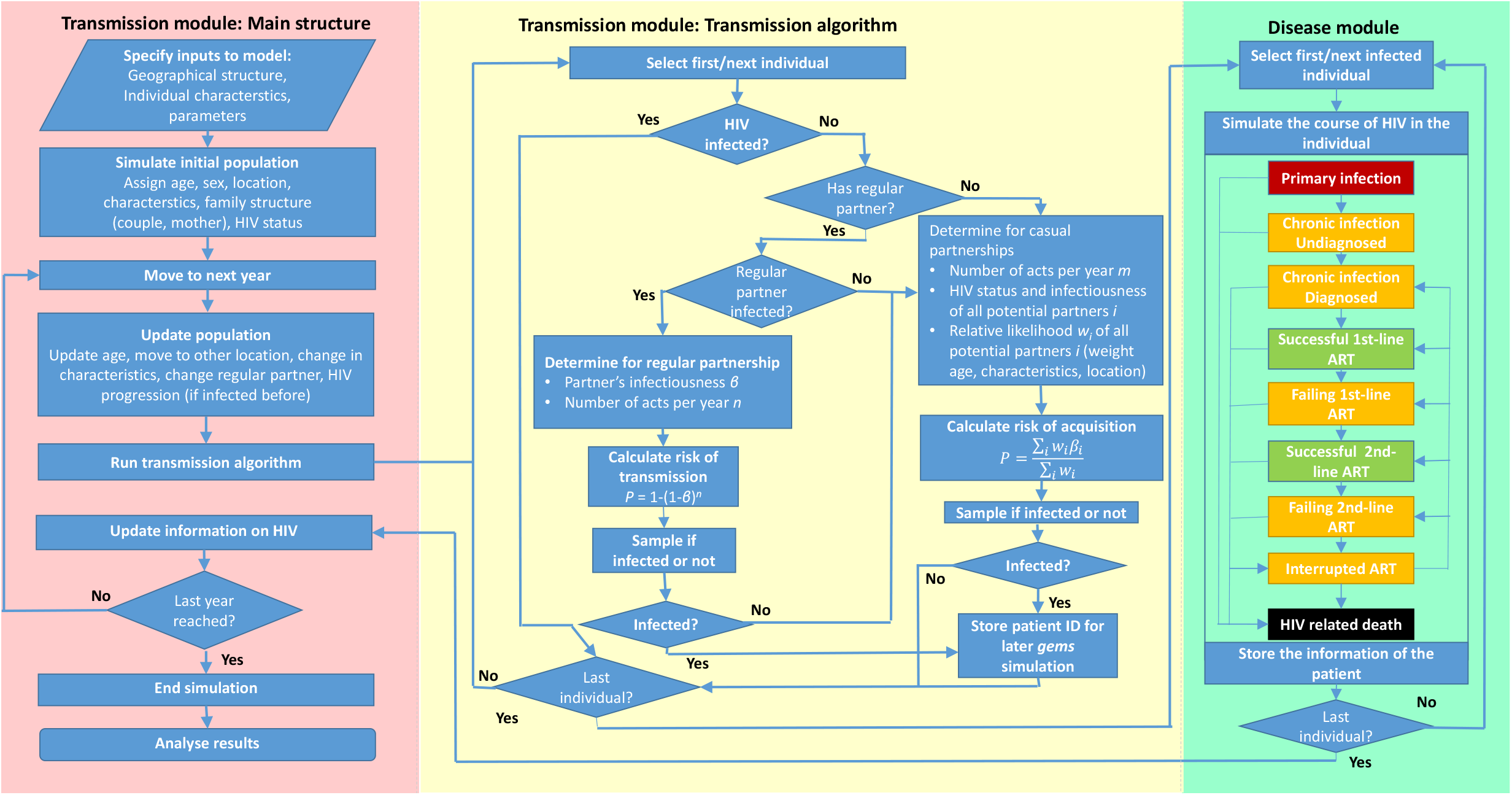
Schematic representation of the mathematical model. The left panel (pink) shows the main structure of the transmission model with a loop over time steps. The middle panel (yellow) shows the transmission algorithm, applied at each time step, in more detail. The right panel (green) shows the HIV disease progression simulation, which is run for each patient at the time of infection.

HIV transmission is determined by an algorithm consisting of two pathways: serodiscordant regular partnerships, or through casual relationships. In serodiscordant partnerships the uninfected partner may acquire HIV, and the risk depends on the infected partner’s HIV status and the frequency of sex acts. Casual partnerships are not modelled explicitly, but an average risk of getting infected is calculated for each individual. The sociobehavioural characteristics determine each individual’s number of casual sex acts, and the characteristics, age and geographical location the likelihood of each potential partner. The number of unprotected sex acts per year in casual and regular partnerships is adjusted with a year-specific coefficient to calibrate the model to observed prevalence estimates.

The *disease module* determines the progression of each infected individual’s HIV infection (Fig. 1; <u>Text S1, Table S2</u>). The module is based on the R package *gems* and is similar to previously developed standalone HIV simulation models.[5-8] The course of infection is divided into health states based on the natural course (primary infection, chronic infection, AIDS, HIV-related death), diagnosis status, and treatment status (off ART, 1^st^ -line or 2^nd^ -line; successful or failing treatment). The timings of transitions between health states are sampled from distributions. Each year, the model is called to simulate the course of infection for all individuals infected in that year. The outputs are then fed back to the transmission module to determine the infectiousness of each individual, and the time of HIV related death.

### Model calibration and scenarios

We simulated the epidemic of Malawi for the years 1975-2030, with the aim to reproduce the differences in the epidemic across districts by the geographical structure alone. To save computing time, preliminary runs were done with a 10% sample of the total Malawian population, and the results were confirmed by running the model for the true population size, adjusting the parameters if necessary. We adapted the prior parameters from a previously published deterministic model, which we adjusted during the calibration of the model.[6] The population was divided into two risk groups that differed in terms of average frequency of unprotected sex acts. We iteratively increased the complexity of the mathematical model by adding geographical diversity, resulting in five alternative models (Table 1; <u>Table S3</u>). At each step, we adjusted the number of annual sex acts in both risk groups by a year-specific coefficient (using the parameters of the previous model as a starting point) so that the national adult HIV prevalence would stay between the UNAIDS lower and upper estimates, or within 5% (relative) margin of these, from 1990 to 2019.

**Table 1.** List of modelled scenarios

| Model | Description | Distinct locations | Movements* between locations | International movements* | Mixing** between locations | Fitted variables |
| --- | --- | --- | --- | --- | --- | --- |
| I | Baseline | 1 | n/a | n/a | n/a | Casual sex acts |
| II | District model | 28 | No | No | No | Casual sex acts, district-level movement* rates |
| III | District model with inter-district transmission | 28 | Yes | No | Yes | Casual sex acts, mixing** rates between districts |
| IV | District model with immigration | 29*** | Yes | Yes | No | Casual sex acts, infection risk abroad |
| V | Grid model with inter-cell transmission | 946 | Yes | No | Yes | Casual sex acts, mixing** rates between cells |

\*Movements refer to permanent relocations (i.e. individuals moving to another geographical location for a duration of at least one year).
\*\*Mixing refers to casual relationships with unprotected sex between individuals who reside in different geographical locations.
\*\*\*The 29<sup>th</sup> location corresponds to the Malawian and Malawi-connected population outside the country, and is treated regarding transmission differently from the remaining locations.

First, we ran the model without any geographical division (Model I). In the next step, we added a geographic dimension by dividing the population into 28 distinct locations, corresponding to the present administrative division of Malawi (Model II). Malawi consists of three regions (Northern, Central and Southern), which in turn are divided into a total of 28 administrative districts. We first assumed that 1% of the population would move to another, randomly selected, district every year, and adjusted the rates of moving to and from certain districts to keep the prevalence close to the census data.[9] We allowed casual sex acts only within each district.

Next, we added the possibility to have casual sex acts between different districts (Model III). We used a distance measure that is based on the minimum number of district borders that need to be crossed between the districts (for example, the distance between neighbouring districts being 1). The likelihood to choose a partner is proportional to the negative exponent of the distance. We compared the district-level prevalence in 2010 with the estimates from DHS.[3,10] We made three attempts to either double or halve the distance scale, in order to either diminish or increase the differences in prevalence between districts, respectively, and get closer to the observed district-level prevalence.

In the following step, we added international migration, using a 29^th^ geographical location representing the population connected to Malawi residing abroad (Model IV). Approximate in- and out-migration rates were taken from the literature, combining estimates on net migration and numbers of migrants living in Malawi.[11] We included at the beginning (1975) an additional population of 2,000,000 individuals who at that time were residing abroad. For people residing each year abroad, the risk of getting infected was determined from the HIV prevalence in the most common destinations of Malawians (South Africa, Zimbabwe, Mozambique). We scaled the risk of infection abroad and the level of casual acts within Malawi to find a balance between domestic and imported infections, leading to desired prevalence levels.

Finally, we ran a model with a finer geographical resolution (Model V). We divided Malawi into 946 cells, corresponding to a 10×10 km^2^ grid. For calculating the district-level prevalence, we determined the districts so that each cell would belong to one district. We set the population size in 1975 for the cells containing the 17 largest densely populated centres of Malawi to be equal to their actual size, and used the average based on each district’s population for the remaining cells. We assumed the same rate of annual permanent moves as in Model II: because of the random allocation of the destination cell, the vast majority of people and households who move would move to another district. For casual relationships, we used mixing based on Euclidean distance, scaled analogously to the distance in Model II (so that the maximum distance within the country would be equivalent). International migration was not considered in this model. We adjusted the sex acts and scale of mixing between cells to calibrate the model for the national and district-level prevalence data.

## Results

In all models, the whole adult population needed at least monthly unprotected sex with a casual partner to reach the observed prevalence level (Fig. 2). The average number of annual casual sex acts producing the best fit in Model I was 20 for low-risk and 70 for high-risk individuals until early 1991. Between 1992 and 1995, the corresponding numbers were set at 16 and 54, between 1996 and 2002 at 10 and 34, and from 2003 onwards, 12 and 40, respectively (<u>Fig. S1</u>). The corresponding numbers in models II to V were similar although small differences existed. HIV prevalence stayed within or close to the UNAIDS estimated range in all models after parameter adjustments. The decreasing trend was projected to continue from 2020 onwards, with the national prevalence reaching about 5% in 2030 in all models.

**Figure 2.**
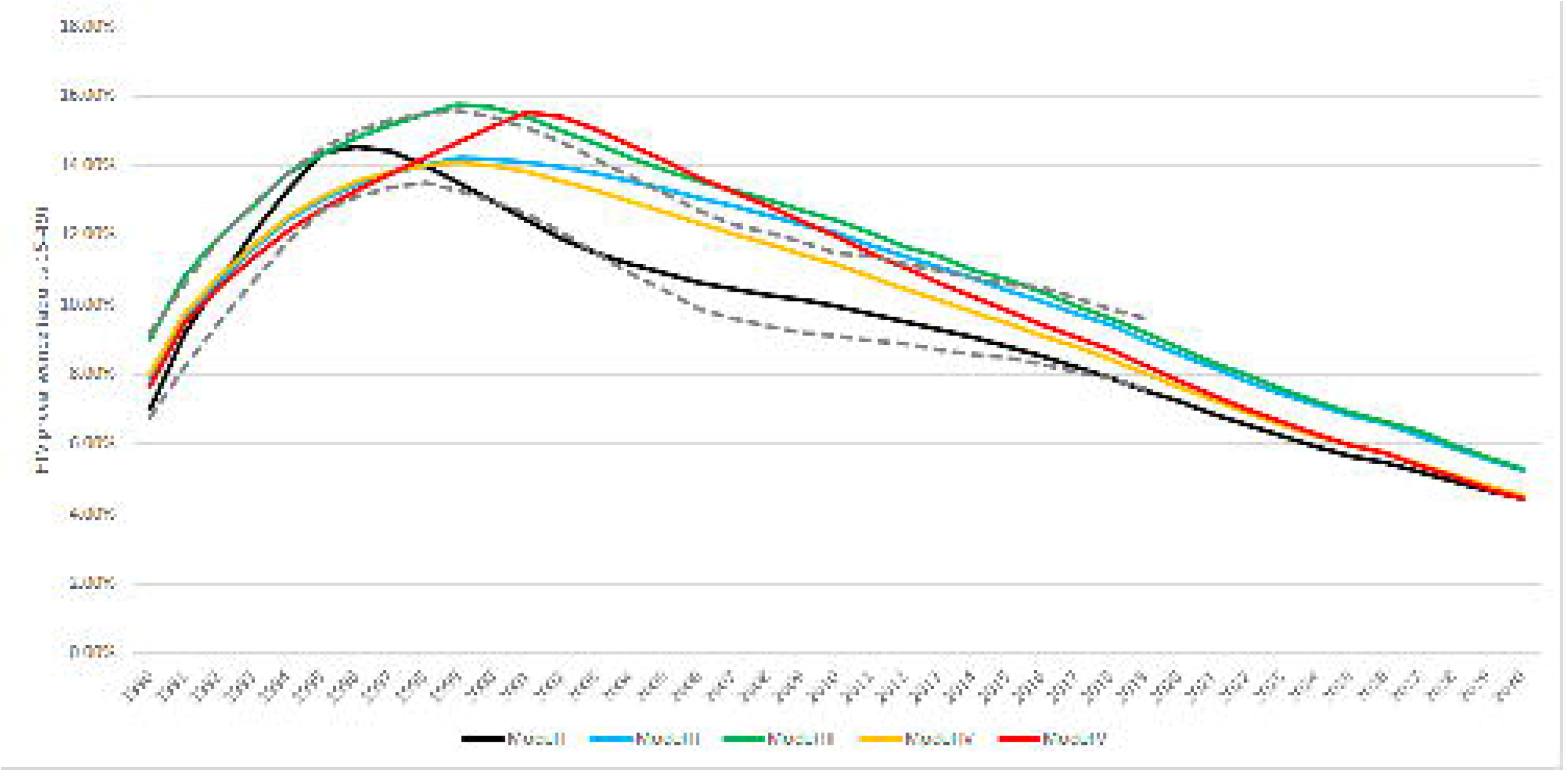
National HIV prevalence among adults aged 15-49 years in Malawi 1990-2030 in the five different models. Model I (black curve): no geographical structure; Model II (blue curve): 28 districts, no international migration, no casual sex between districts; Model III (green curve): 28 districts, casual sex between districts allowed, no international migration; Model IV (yellow curve): 28 districts, casual sex between districts allowed, international migration considered; Model V (red curve): 946 10×10km^2^ cells, casual sex between cells allowed, no international migration. The dashed curves in panel A show the UNAIDS projections.

In the models with districts (Models II to IV), the moving rates out of Dedza, Dowa, Ntcheu and Zomba districts had to be increased from the default level 1% to 2% per year, and out of all other districts except Chitipa, Karonga, Mzimba, Lilongwe, Machinga, Mangochi and Neno to a lesser extent, to keep within the population distribution (<u>Supplemental Digital Content Table S4</u>). In turn, the attractivity of Neno district as a destination had to be increased by 50%. With these assumptions, the population sizes of all districts remained within a 10% margin of the census data.

When Likoma Island (no data) is excluded, the observed district-level prevalence among adults aged 15-49 years in 2010 ranged between 4.4% (Chitipa) and 21.6% (Thyolo), the prevalence being generally highest in the Southern region.[10] The heterogeneity between the three regions was preserved in the district-level model without international migration or inter-district causal relationships (Model II; Fig. 3). The prevalence distribution in 2010 was highest in the same districts where the infections were seeded in 1975, and the prevalence was generally higher in the Southern region than the rest of the country. The models however could not reproduce the finer-scale patterns, such as the concentration of highest prevalence into the densely populated districts of Chiradzulu, Mulanje and Thyolo, or the zone of notably low prevalence on the north side of the capital Lilongwe. The modelled prevalence in 2010 was highest in Balaka (16.2%) and Nsanje (16.0%) districts, and clearly lowest in Likoma (2.8%), followed by Dowa (7.3%). In 2030, the highest prevalence was projected in Nsanje (6.4%).

**Figure 3.**
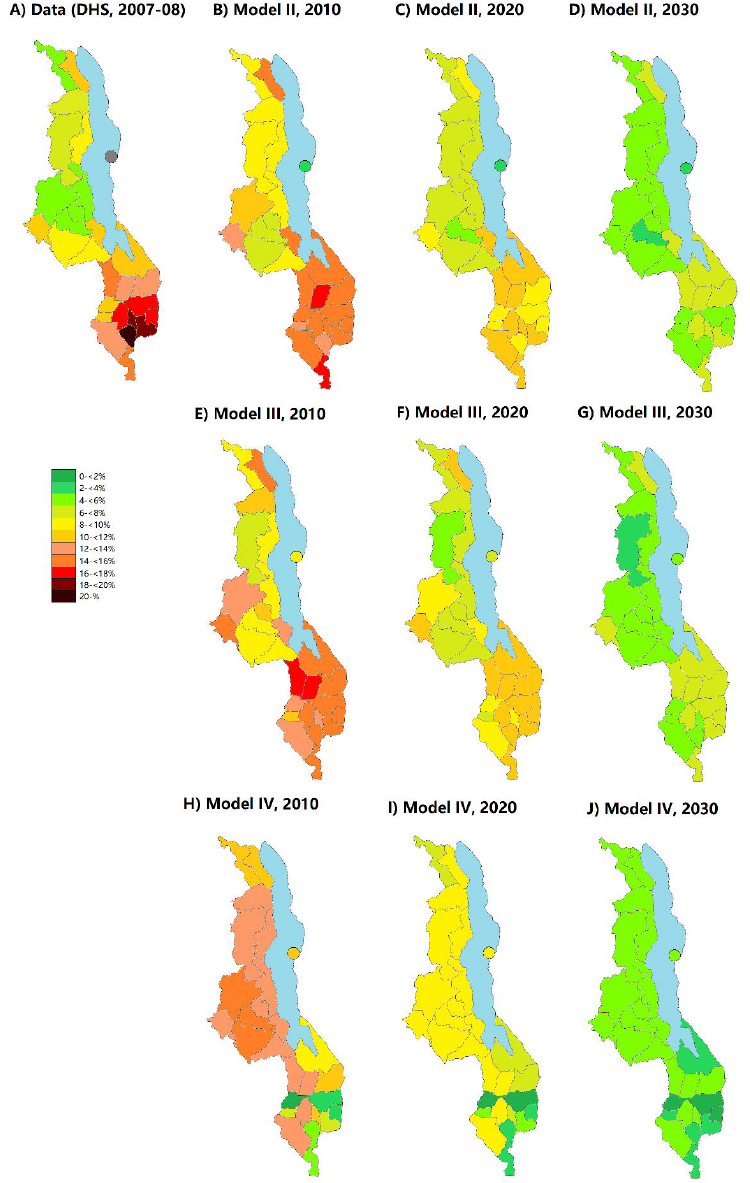
HIV prevalence among adults aged 15-49 years in 2010, 2020 and 2030 in the 28 administrative districts of Malawi in models with district-level geographical structure. Panel A: Data from the Demographic and Health Surveys (DHS) in 2010. Panels B-D: Model II (no international migration, no casual sex between districts). Panels E-G: Model III (casual sex between districts allowed; no international migration). Panels H-J: Model IV (casual sex between districts allowed; international migration included).

In Model III, the differences in HIV prevalence between districts almost disappeared in the first version where the distance between neighbouring districts was set to one unit. By doubling the distance three times (to 8 units per crossed district border), district-level prevalence in 2010 became clear. This metric can be interpreted as a person choosing a casual partner *e*^8^ = 2980 times more likely from his own district than from a neighbouring district. Similar to Model II, the prevalence in 2010 was highest in the Southern region and in the few other districts where infections were seeded at the beginning of the model, but the observed heterogeneity between individual districts could not be reproduced (Fig.3). The lowest modelled prevalence in 2010 was in Mzimba (6.9%) and the highest in Balaka (16.9%). In 2030, the range across districts was 4.0%-6.3%.

In contrast to Models II and III, in Model IV the geographical pattern disappeared: the prevalence in 2010 was highest in the central region (about 14% in Lilongwe and the surrounding districts). The lowest prevalences were projected in Neno (1.9%), Zomba (2.4%) and Phalombe (3.3%), the two latter being known to have prevalence estimates above 16% in reality. In Model IV, the risk of getting infected had to be made twice as high than what would be expected by the prevalence abroad in order to keep the national prevalence on an acceptable level. In 2030, the projected prevalence was below 6% in all districts in all three models.

In Model V with a high geographical resolution, the geographical heterogeneity across districts in year 2010 was approximately in line with Models II and III, ranging from 6.0% (Nkhata Bay) to 16.0% (Phalombe; Fig. 4). Small clusters of cells with prevalence around 20% were found across the Southern region, but also elsewhere, for example in Mchinji district, and along the north and south borders of Rumphi district in the Northern region. The evolution of the district-level prevalence over time was also similar to Models II and III. In 2030, the prevalence was above 6% in some individual cells, mainly in the same cells that had the highest prevalence already in 2010; none of the cells were HIV free.

**Figure 4.**
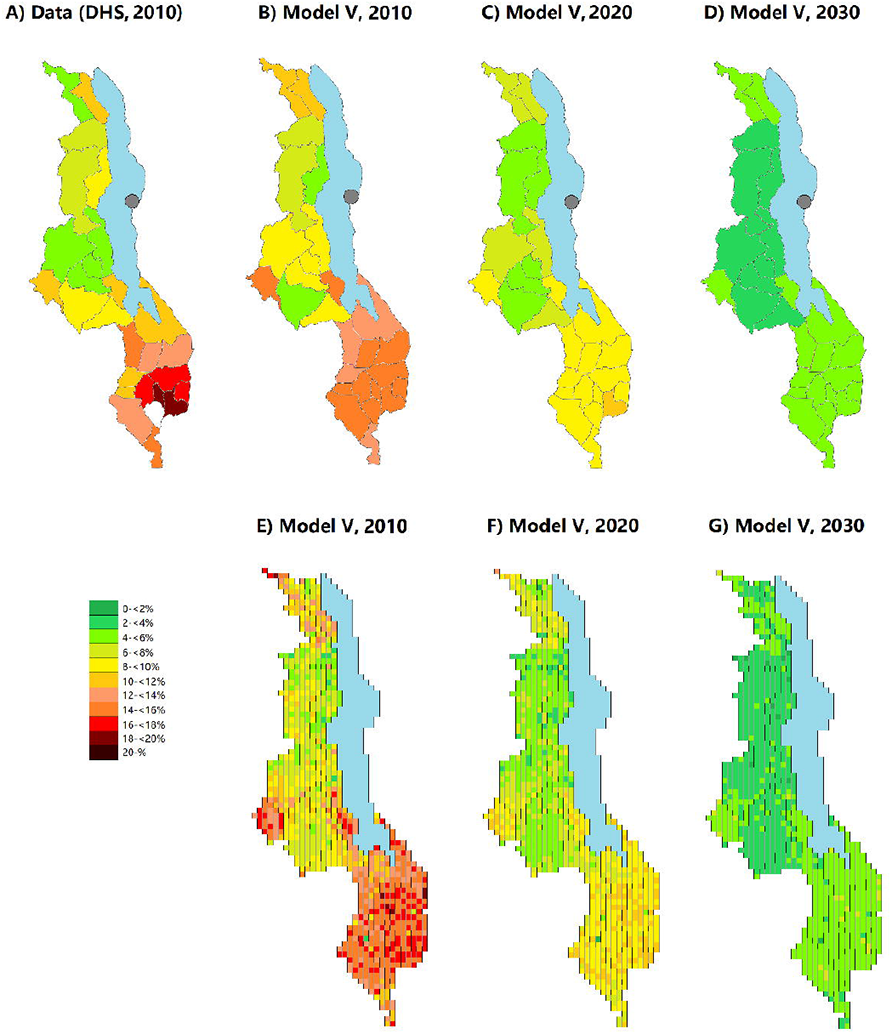
HIV prevalence among adults aged 15-49 years in 2010, 2020 and 2030 in Malawi in the model with a geographical resolution of 10×10 km^2^ (Model V). Panel A: Data from the Demographic and Health Surveys (DHS) in 2010 for the 28 administrative districts. Panels B-D: Model estimates for the 28 administrative districts. Panels E-G: Model estimates for the 946 cells.

## Discussion

A model with simple geographical representation can reproduce the prevalence patterns observed across Malawi to some extent. The differences in later years were essentially a result of the initial distribution of the infections at the beginning of the simulation in year 1975. A substantial mixing between people residing in different areas can smoothen the prevalence across the country very fast: the heterogeneity could be kept only if sexual relationships between people living in different geographical locations were restricted to a minimum. Considering international mobility within a reasonable rate diminished the differences in HIV prevalence rapidly. Increasing the geographical resolution of the model did not essentially influence the district-level prevalence estimates.

We started all simulations with the assumption that HIV was present in 1975 only in districts where the prevalence in 2010 was at least 10%. This includes all districts of the Southern region, as well as the districts Mchinji, Ntcheu and Salima (Central region) and Karonga (Northern region). This assumption is somewhat arbitrary. The first case of HIV was officially detected in Malawi in 1985, but it is reasonable to assume that the disease had been spreading in the country already years before.[12] The origin of HIV-1 has been localized to Central Africa.[13] Whether HIV had already spread to the entire Malawi or only selected locations by mid-1970s is unknown. A seroprevalence survey from Uganda from early 1970s found widespread antibodies in the population, suggesting that the infection was widespread in Africa already at that time.[14] But as rural areas tend to be overall less affected by HIV, it could be likely that HIV was introduced only later in places like North Malawi, which are less densely populated and have limited connections to other regions.[15] Moreover, there was a strong movement in the early 1970s of Malawian migrant workers moving back to the country from the neighbouring countries, mainly to take jobs in the growing agricultural export sector.[16] Most large plantations are located in the Southern region, so it could well be that the differences in prevalence date back to the 1970s, supporting the assumption that the spread of HIV in Malawi started in the South or other particular areas. In turn, when we explicitly included international migration from 1975 onwards in the model we could no longer reproduce the observed geographical heterogeneity. It may be that migration indeed plays a lesser role in the Malawian HIV epidemic in the recent decades. On the other hand, this shows that the assumptions on migration can entirely change the model’s results, so more attention should be paid on the true role of international mobility on the HIV epidemic.

The fitted parameter for sex acts was notably high: the observed high prevalence could be reached only if all individuals had sex outside the regular partnerships at least once every three weeks, which is hardly realistic. This raises questions about the need of additional features into the model that would enhance the heterogeneity of partnerships structures. First, the high-risk population could be further categorized to include also “superspreaders”.[17] Second, the structure of regular partnerships may need to be diversified. In Malawi, 15% of men and 27% of women are estimated to be living in polygamous relationships:[18] allowing polygamous relationships with partial concurrence in the model may thus be more realistic than the current approach restricted to distinct or sequential monogamous partnerships. Third, male-to-male partnerships, currently excluded, may accelerate the spread of HIV. The biological risk of transmission per act is about 10 times higher between males than males to females;[19] and because homosexuality in Malawi remains illegal and highly stigmatized, most men having sex with men are likely to also have regular or casual female partners.[20]

All five parameterisations confirmed the future decreasing trend of HIV prevalence: we expect that by 2030, the national prevalence will be around 5%. The differences between districts will also even out. While in 2010, the model’s outputs were roughly in line with the DHS estimates, as of now (2020) the range was projected at 3% to 10%, and by 2030 no district would have a prevalence higher than 6%. Data from the Malawi Population-based HIV Impact Assessment (MPHIA) showed that differences across geographical districts still existed in 2015, and the trends were similar as in the 2010 DHS survey.[21] The differences may thus be levelling out slower than according to our models. This would support the hypothesis that the differences do not only depend on transmission dynamics but also on risk behaviour, which in turn could be influenced by sociobehavioural factors. Direct determinants of the risk of acquiring HIV, such as the number of unprotected sex acts and variability of partners, are strongly associated with social determinants. In the complex network of factors potentially associated with HIV, urbanity and literacy were the most central variables.

The model with fine resolution led to similar results as the district-level models, with a few characteristics worth noting. The smoothening of prevalence between districts was faster than within the district-level models particularly in the Northern region, despite the fact that in only one of the six Northern districts (Karonga) HIV was assumed to be present in 1975. This could be due to the population density: in the model with fine resolution, in the North each cell had a much smaller population than the South, meaning that more people had to seek partners from outside their own cell. In turn, in the district model the number of people per district was relatively similar, since the districts in the North tend to have larger areas than in the South. Another interesting pattern was the prevalence patterns in the border areas and along the lakeshore. For example, the narrow strip in Mangochi District between Lake Malawi and Mozambique border had cells with both very high and very low prevalence. These cells have only few other cells within a small radius, so people tend to seek partners more frequently from their own cell than in inland locations far from the borders. A similar pattern was seen in the district of Nsanje, which is almost completely surrounded by national border. Nsanje had a much lower prevalence than the rest of the Southern region in the fine resolution model, which contradicts both the data and the other models. In regions with few connections between cells, chance is likely to play also a major role in how the epidemic will develop.

Our study had several limitations. Because of the computationally expensive model structure, the parameter fitting was done on an ad hoc basis, and parameter uncertainty or the stochastic variability of the results was not estimated. The model’s findings were also based on the arbitrarily chosen distribution of HIV in 1975. Several key factors were ignored in the model, including polygamy and concurrent regular partnerships, male-to-male transmission, and the associations between high-risk behaviour and mobility. However, the model developed for this study serves as a basis for further extensions.

## Conclusions

The high prevalence in Southern Malawi may have developed partly as a result of an earlier introduction of HIV into this region. This could possibly be related to the return of Malawians from abroad in the 1970s. On the other hand, the models required a high number of sexual contacts for the entire population to realistically represent the further spread of the epidemic. The obvious explanation would be that there are also behavioural factors that accelerated transmission, particularly in the Southern region. Our results of this project form a basis for further evaluation and understanding of both the Malawian and global HIV epidemic: our model can take into consideration, in addition to the geographical dimension, an arbitrary number of individual-level factors, and can easily be adapted to other countries or settings. Whereas the broad difference between Southern Malawi and the rest of the country may well be due to the connection network between districts, the variability of the socio-behavioural factors and their impact on HIV transmission needs to be quantified. Implementing these differences into mathematical models may help to get an in-depth understanding of the differences in HIV prevalence within countries.

## Data Availability

The study uses only publicly available data.

## Acknowledgments

OK was supported by the Swiss National Science Foundation (grant 163878).

## Supporting information

Supporting information.docx (contains the following items):

**Text S1. Technical details of the model**

**Table S1. Parameters of the transmission model**.

**Table S2. Parameters of the disease progression model. Table S3. Parameters related to the geographical dimension**.

**Table S4. District-specific in- and out-movement rates (models II to V)**.

**Figure S1. Average annual unprotected sex acts with casual partners in the models I to V**.

