## Supporting Information for "The spatial spread of HIV in Malawi: An individual-based mathematical model"

Janne ESTILL, Wingston NG’AMBI, Liudmila ROZANOVA, Olivia KEISER

Contents:

Text S1. Technical details of the model

Table S1. Parameters of the transmission model.

Table S2. Parameters of the disease progression model.

Table S3. Parameters related to the geographical dimension.

Table S4. District-specific in- and out-movement rates (models II to V).

Figure S1. Average annual unprotected sex acts with casual partners in the models I to V.

References

**Text S1. Technical details of the model**

The mathematical model we developed is a stochastic, individual based model of HIV transmission. The model consists of two modules: a *transmission module*, which is used to update and track the demographics, characteristics, HIV status and geographical distribution of the population; and a *disease module*, which simulates the course of the HIV infection within individual patients.

The model code is available on GitLab (<https://gitlab.com/igh-idmm-public/agent-based-hiv-transmission-model>).

1. Transmission module

The transmission module is represented as a *n*x12-matrix, where *n* is the total number of simulated individuals who ever have been alive since the initiation of the simulation. The columns represent the following variables: (1) individual ID (running number); (2) current age (in years); (3) sex (male, female); (4) current place of residence (code of geographical location); (5) sociobehavioural characteristics (combined code); (6) biomedical characteristics (combined code); (7) HIV status (infected or uninfected); (8) current stage of HIV infection (health state at disease module at the start of year); (9) year of HIV infection; (10) year of death (if died; otherwise zero); (11) ID of regular partner; (12) ID of mother. The model is updated in time step of one year. At each time step, several functions are applied to update the table; newborn individuals are attached to the bottom of the matrix as new rows. Before moving to the next time step, key indicators are collected and stored for later analysis.

*1.1 Creating the initial population*

At the beginning of the simulation, an initial population of a given size is created. The population is assigned demographic characteristics (age, sex, location, sociobehavioural and biomedical characteristics). After this, a given proportion of individuals aged 15 and above are assigned a partner. For partner selection, the same function is used as later to determine new partners after partner change (see details in Section 1.2.3). For individuals aged below 15, a mother (woman who was in childbearing age at the time of the child’s birth and lives in the same geographical location) is also randomly assigned.

A random sample of individuals are then determined to be HIV infected (prevalence determined by sex, age, location and sociobehavioural profile), and for each HIV infected individual a uniformly distributed time of infection within the past five years is assigned. After this, the disease module is run for each infected individual (see Section 2 below).

*1.2 Updates in each time step*

At each time step, the model’s population is updated in the following ways. In the present project, we update the model in time steps of one year, but the updating frequency can be chosen arbitrarily. All parameters and variables related to time need to be defined using the time step as unit (the results can be converted to years, days or other desired unit after the simulation).

1.2.1 Age: Age is updated in the natural way, by adding 1 to the age variable.

1.2.2 Place of residence: In models with a geographical structure (i.e. more than one distinct non-empty location), a proportion of individuals will change location at every time step. The relocation function is applied to all men aged 15 or above, and women aged 15 or above who have no regular partner. Children will follow their mothers, and women in regular partnerships their partners. The relocation process is sampled in two steps. First, the overall probability of moving out is determined by multiplying an overall (constant) annual relocation probability with coefficients depending on age, current location and year. For those who are determined to move, the destination will be chosen from a distance matrix, which determines the relative likelihood of choosing each destination, as well as the present population size of the destination location. The relative probabilities of each destination are determined by multiplying the population size of each district with the row of the distance matrix that corresponds to the origin location, as well as with a year- and destination-specific multiplier that takes into account fluctuations over time in the attractivity of each district. The last location represents “abroad”, i.e. the population with connections to the modelled setting who at present are residing somewhere else. Technically, “abroad” is treated in a similar way to any other location.

1.2.3 Forming regular partnership: For each person aged 15 or above, the model will decide if the person will deform or form a partnership. First, the model checks the current partnership status. Those currently in partnership will split up according to a fixed probability; the partnership status of them and their partners will be set to having no partner. Second, the model checks who of those currently having no partner wish to start a new partnership. The selection is done for each geographical location separately, assuming that new partners are always chosen from the same location. Within a location, all men and women waiting for partners are listed. Then, the likelihood of different partners is calculated using two mixing matrices: for age and sociobehavioural characteristics. This is done as a loop of all combinations of age group and sociobehavioural characteristics for males. Males with this combination of age and characteristics will choose partners from all women, whose probabilities are weighted based on the mixing matrices. If there are overall more men than women looking for partners, the option to be left without a partner is also included in the selection process; if the opposite, those women who are left last will remain without partners.

1.2.4 Reassigning sociobehavioural characteristics: Sociobehavioural characteristics are represented as one integer variable, which can be decoded into different indicators (for example, if there were three included variables with 2, 4 and 10 values each, the combined variable would have values between 0 and 79 (80=2*4*10 categories). In the present project, we however only include one variable with two possible values (low or high risk). There is a pre-defined matrix for transitions between the sociobehavioural categories, the element *i*,*j* representing the conditional probability that an individual in category *i* in the previous year will be in category *j* in the present year.

1.2.5 Reassigning biomedical charactristics: The biomedical characteristics are presented and updated in the same way as the sociobehavioural characteristics. In the present version of the model, specific biomedical characteristics are not included.

1.2.6 HIV transmission: HIV transmission is modelled in two steps, sampling infections in regular and casual partnerships, respectively. For any contact (single unprotected sex act) between an infected and uninfected individual, the probability of transmission depends on the stage of HIV infection the infected partner is in (primary, chronic untreated, virologically suppressed, on failing treatment, interrupted treatment), the types of sex (female to male, male to female, male to male), and biomedical characteristics that influence the risk of transmission. Transmissions in regular partnerships are determined by calculating the annual risk of transmission from the annual sex acts and per-act probability (as described above) for each serodiscordant partnership. Casual partnerships in turn are not explicitly modelled, instead the model calculates for each person a risk of getting infected based on the current prevalence of HIV among the expected partners. This is done in the following steps. First, each person is assigned a number of unprotected heterosexual acts per year depending on his/her sociobehavioural category. The acts are then balanced so that men and women have the same total number. Second, in order to make the model computationally efficient, the individuals are split into groups according to their location, age (categorized) and sociobehavioural and biomedical characteristics. Each person is thus assigned an indicator that tells exactly the values of all variables mentioned before. Then, in a loop over all categories, the model calculates for the women in the category the sum of total sex acts by men over each category, weighted by the relative probabilities of contact between the categories (which in turn depends on the mixing matrices between different locations, age groups, and sociobehavioural and biomedical categories). Then, a similar sum is calculated, but only considering the HIV infected males and multiplying each potential act by the infectiousness. An average risk of infection per act is then gained by dividing the former with the latter. Next, the same process is repeated for men to calculate the potential acts and infectiousness of female partners. Finally, the risk of infection per act for individuals residing currently abroad is calculated from the prevalence abroad. Afterwards, a similar process is repeated for men having sex with men (i.e. male persons with a sociobehavioural characteristic category representing preference for male-to-male sex; not applicable in the current version of the model).

Finally, the risk to get infected is calculated for each uninfected individual using the per-act transmission risk calculated as mentioned before, and the number of sex acts. For each uninfected individual, the model samples if he/she will get infected during the year, based on this calculated probability. Once the newly infected individuals are identified, the HIV simulation module is run to determine the course of their infection. The time of infection is also recorded for the newly infected individuals.

1.2.7 Updating the HIV status: Each year, the status of HIV infected patients at the end of the year is recorded from the outputs of the disease progression module.

1.2.8 Births: The births of new infants is modelled by sampling from fertility rates that are determined for each woman based on age (5-year age groups) and the year. After determining the births, each child is given a time of birth (randomly distributed during the year), sex (equal probabilities), location (location of the mother), sociobehavioural and biomedical characteristics (in the current version, the default values), and HIV infection. HIV infection is sampled for infants born to HIV infected mothers from probabilities depending on the mother’s HIV status and year (to account for the availability of PMTCT services). For infants determined to be HIV infected, the disease progression simulation is run, just like for adults.

1.2.9 Deaths: Deaths can happen from two reasons which as modelled separately. Deaths not related to HIV are sampled from HIV-free age- and sex-specific mortality rates (i.e. annual probabilities). HIV related deaths are determined already in the disease progression simulation: each year, the transmission model checks from the results of the simulation which HIV infected individuals proceed to the last stage of the HIV progression (death) and assigns these as died. The rows for died people will remain in the matrix, but no longer updated.

*1.3 Collecting outputs*

The basic output of the model is the population matrix. Since the matrix is updated for every year and the previous data are overwritten, some indicators cannot be derived from this in real time (for example, the location in the final version corresponds to the location of the individual at the last year of simulation or his death). Therefore, the model calls after each year an output collector script, which stores the results of interest at the end of each year.

2. Disease progression module

The disease progression module is a separate model, which can be used both as a part of the transmission model, or as a standalone simulator of HIV cohorts. The module is realized using *gems*, an R package for generalized multistate simulations.[1] The functioning of *gems* is described in detail elsewhere: in brief, the progression of the disease is represented as a directed acyclic graph of health states and transitions between them; each allowed transition is assigned a hazard function that may depend on time, the individual’s characteristics, and the history of previous events. Once these are determined, a cohort of individuals is simulated. Each individual starts in the initial heath state. Transition times for all possible transitions are sampled from the corresponding hazard functions; the smallest of these times determines to which state the individual moves and when. This process is then repeated from the current state, until the individual either reaches a terminal state, or the maximum follow-up time; and the model then moves to the next individual. The output of the simulation is the set of entry times to each state for each simulated individual.

In this model, the progression of HIV is divided into 18 states: primary infection (1); undiagnosed chronic infection (2); diagnosed chronic infection (3); successful first-line treatment (4,9,14); failing first-line treatment (5,10,15); successful second-line treatment (6,11,16); failing second-line treatment (7,12,17); interrupted treatment (8,13); and HIV-related death (18). The structure allows up to two interruptions of treatment (because *gems* does not support return back to a previously visited state, each stage of treatment needs to be represented with three separate states). For each possible transition, a function (either time-depending hazard, or fixed time to event) is defined. The only baseline characteristic that we currently need is the calendar time of infection. Primary infection is defined to last for 3 months; diagnosis is assumed to be possible from year 1990 onwards according to a year-depending rate (newborn infants, and women of childbearing age from 2011 onwards, have differing rates). Rates of treatment failure, switching therapy, interrupting treatment and returning back to care are adapted from our previous studies. When returning to care after interruption, the patient will go to the stage of treatment he/she was before the interruption.

3. Additional details for fitting the models

*3.1 Model I (baseline model)*

We fitted the annual number of unprotected sex acts with casual partners. Initially we started with the default assumption of 3 acts per year for low-risk individuals, and 10 acts per year for high-risk individuals, regardless of sex. These numbers were then multiplied by yearly coefficients, so that the prevalence would remain within the UNAIDS estimates from 1990 onwards.[2] By allowing a yearly adjustment, we took into account the adaptive behaviour, for example because of increased awareness of HIV. The results of this fitting were taken as a starting point for models II-V as well, and adjusted further if necessary.

*3.2 Model II*

The rate of relocation between districts was fitted to the observed population estimates.[3] We first assumed that 1% of all households would relocate each year, with a random allocation of the destination district. If the population of a district grew too fast, we increased the rate of moving out; and if the population grew too slowly, we increased the weight of the corresponding district as a destination.

*3.3 Model III*

We used a metric where the distance between two districts depends on the minimum number of borders that need to be crossed in between, i.e. that the distance of a district to itself is 0, to a neighbouring district 1, to a district with common neighbour 2, etc. In the initial model, the likelihood of having a partner was assumed to correlate inversely with the exponent of the distance, i.e. when choosing a partner for casual relationship, each potential partner is given a weight *e*^-^*^d^*^(^*^i^*^)^ where *d*(*i*) is the distance from the partner’s district to the index person’s district. After the simulation, we compared the prevalence in each district to the 2010 DHS estimates.[4] If the differences between districts were too small, we doubled the distance between districts (i.e. multiplied the original distance measure by two); if the differences were too large, we halved the differences. We did a maximum of three iterations.

*3.4 Model IV*

In Model IV, we explored the role of international migration. As a baseline assumption, we used the average prevalence in South Africa, Zimbabwe and Mozambique to determine the risk of acquiring HIV for people living abroad.[2]

*3.5 Model V*

Model V used a 10x10 km^2^ grid, dividing Malawi into a total of 946 square-shaped cells. The parameterisation was done in line with Model III. For permanent relocation, the probability of choosing the destination location was made random; because of this, we kept the annual rate of moving the same as in Model III, as only a small proportion of those who move would choose a destination from the same administrative district. The distance for transmission probability was based on Euclidean distance (calculated the standard way i.e. square root of the sum of squares of longitudinal and latitudinal coordinates), scaled so that the maximal distance between cells (i.e. the northernmost and southernmost tips of Malawi) would be the same as the maximum distance in Model III.

**Table S1. Parameters of the transmission model.**

| **Parameter** | **Value** | **Source** |
| --- | --- | --- |
| ***Initial conditions in 1975*** |  |  |
| Total population size | 5 302 000 | [3] |
| Proportion of women | 50.1% | [3] |
| Age distribution | See reference | [3] |
| HIV infected population | 4000 | Assumption, [5] |
| Proportion of women with high-risk behaviour | 5% | Assumption |
| Proportion of men who are high-risk | 5% | Assumption |
| Proportion of HIV infected who are women | 50% | Assumption |
| Proportion of HIV infected who are high-risk | 50% | Assumption |
| ***Yearly progression*** |  |  |
| Probability to move from high to low risk: women | 0.04/year | Assumption, [5] |
| Probability to move from high to low risk: men | 0.10/year | Assumption, [5] |
| HIV-free mortality: children aged 0-14 | 0.014/year | [6] |
| HIV-free mortality: women aged 15-49 | 0.005/year | [6] |
| HIV-free mortality: men aged 15-49 | 0.006/year | [6] |
| HIV-free mortality: women aged 50 or above | 0.042/year | [6] |
| HIV-free mortality: men aged 50 or above | 0.048/year | [6] |
| Birth rate | 0.209/year | [5,7] |
| Probability of change of regular partners (until age 50) | 1/year | Assumption, [5] |
| Proportion of homogeneous mixing for regular partnerships | 100% | Assumption |
| Proportion of homogeneous mixing for casual partnerships | 100% | Assumption |
| Per-act HIV infectiousness male to female, chronic phase | 0.0019/act | [8] |
| Per-act HIV infectiousness female to male, chronic phase | 0.0010/act | [8] |
| Relative infectiousness in acute phase* | 20 | [9] |
| Relative infectiousness while treated (successful treatment)* | 0.07 | [2] |
| Relative infectiousness while on failing treatment * | 0.40 | Assumption |
| Mean number of unprotected sex acts with regular partner | 50/year | Assumption |

*Compared with chronic phase

**Table S2. Parameters of the disease progression model.**

| First year HIV can be diagnosed | 1990 | Assumption |
| --- | --- | --- |
| First year ART is available | 2003 | [10] |
| First year PMTCT is universally available | 2011 | [10] |
| First year ART is broadly available | 2011 | [10] |
| First year virological monitoring on ART is available universally | 2012 | [10] |
| First year ART is available immediately after diagnosis | 2020 | [11] |
| Diagnosis rate (except for infants and women in PMTCT) | 0.05/year | Assumption |
| Diagnosis rate (women in PMTCT) | 1.00/year | Assumption |
| ART initiation rate before broad availability of ART | 0.07/year | Assumption |
| ART initiation rate after broad availability of ART | 0.40/year | Assumption |
| ART initiation rate after universal availability of ART | 1.00/year | Assumption |
| Treatment failure rate | 0.05/year | Assumption,[5] |
| Switching rate without treatment failure without virol. monitoring | 0.01/year | Assumption,[5] |
| Switching rate with true treatment failure without virological monitoring | 0.20/year | Assumption,[5] |
| Switching rate with true treatment failure and virological monitoring | 1.00/year | Assumption |
| Dropout rate in the first year of treatment | 0.25/year | Assumption,[5] |
| Dropout rate from the second year of treatment onwards | 0.10/year | Assumption,[5] |
| Rate of returning back to care | 0.33/year | Assumption,[5] |
| Duration of acute phase | 3 months | [9] |
| Mortality during chronic phase | 0.05/year | Assumption,[5] |
| Mortality on successful ART | 0.01/year | Assumption,[5] |
| Mortality on failing ART | 0.10/year | Assumption,[5] |

**Table S3. Parameters related to the geographical dimension.** Models II-IV refer to the models with district-level resolution, Model V to the model with 10x10 km^2^ resolution.

| **District** | **Models II-IV** | | | **Model V** | | |
| --- | --- | --- | --- | --- | --- | --- |
|  | Population (1975) | Neighbouring districts | International border | Number of cells | Population per 10x10km^2^ cell excluding cities* (1975) | Cities* and their population (1975) |
| Chitipa | 63,600 | Karonga, Rumphi | Yes | 43 | 1700 | - |
| Karonga | 95,400 | Chitipa, Rumphi | Yes | 34 | 2800 | Karonga 14700 |
| Likoma | 5,300 | - | No | 0 | - | - |
| Mzimba | 302,200 | Nkhata Bay, Rumphi, Kasungu, Nkhotakota | Yes | 106 | 2600 | Mzuzu 18700  Mzimba 7600 |
| Nkhata Bay | 84,800 | Mzimba, Rumphi, Nkhotakota | No | 42 | 2300 | - |
| Rumphi | 63,600 | Chitipa, Karonga, Mzimba, Nkhata Bay | Yes | 46 | 1300 | Rumphi 5300 |
| Dedza | 312,800 | Lilongwe, Ntcheu, Salima, Mangochi | Yes | 38 | 7700 | Dedza 13100 |
| Dowa | 281,000 | Kasungu, Lilongwe, Ntchisi, Salima | No | 31 | 8000 | - |
| Kasungu | 238,600 | Mzimba, Dowa, Lilongwe, Mchinji, Nkhotakota, Ntchisi | Yes | 80 | 2300 | Kasungu 8800 |
| Lilongwe | 673,400 | Dedza, Dowa, Kasungu, Mchinji, Salima | Yes | 62 | 9800 | Lilongwe 108500 |
| Mchinji | 164,400 | Kasungu, Lilongwe | Yes | 31 | 5100 | Mchinji 7100 |
| Nkhotakota | 116,600 | Mzimba, Nkhata Bay, Kasungu, Ntchisi, Salima | No | 43 | 2000 | Nkhotakota 12300 |
| Ntcheu | 238,600 | Dedza, Balaka, Mangochi, Neno | Yes | 33 | 6100 | - |
| Ntchisi | 100,700 | Dowa, Kasungu, Nkhotakota, Salima | No | 17 | 5100 | - |
| Salima | 121,900 | Dedza, Dowa, Lilongwe, Nkhotakota, Ntchisi | No | 22 | 5800 | Salima 10500 |
| Balaka | 153,800 | Ntcheu, Machinga, Mangochi, Zomba, Neno | No | 21 | 6700 | Balaka 12300 |
| Blantyre | 403,000 | Chikwawa, Chiradzulu, Thyolo, Zomba, Neno | No | 20 | 9300 | Blantyre 231400 |
| Chikwawa | 180,300 | Blantyre, Mwanza, Nsanje, Thyolo, Neno | Yes | 49 | 4000 | - |
| Chiradzulu | 116,600 | Blantyre, Mulanje, Thyolo, Zomba | No | 8 | 22000 | - |
| Machinga | 185,600 | Balaka, Mangochi, Zomba | Yes | 36 | 5300 | Machinga 9000 |
| Mangochi | 307,600 | Dedza, Ntcheu, Balaka, Machinga | Yes | 67 | 4500 | Mangochi 7800 |
| Mulanje | 212,100 | Chiradzulu, Phalombe, Thyolo, Zomba | Yes | 20 | 15200 | Mulanje 18200 |
| Mwanza | 42,400 | Chikwawa, Neno | Yes | 8 | 4100 | - |
| Nsanje | 95,400 | Chikwawa, Thyolo | Yes | 19 | 5400 | Nsanje 11500 |
| Thyolo | 228,000 | Blantyre, Chikwawa, Chiradzulu, Mulanje, Nsanje | Yes | 17 | 18900 | - |
| Phalombe | 116,600 | Mulanje, Zomba | Yes | 13 | 13100 | - |
| Zomba | 371,100 | Balaka, Blantyre, Chiradzulu, Machinga, Mulanje, Phalombe | Yes | 24 | 13700 | Zomba 37900 |
| Neno | 26,500 | Ntcheu, Balaka, Blantyre, Chikwawa, Mwanza | Yes | 16 | 2400 | - |

*”Cities” refer to densely populated national or regional centres, not necessarily corresponding to the formal definition of a city.

**Table S4. District-specific in- and out-movement rates (models II to V).** The probabilities to choose a specific destination district are determined by multiplying the population size with the coefficient in the in-migration column below, and normalizing with the sum over all districts.

| **District** | Out-migration (proportion moving out every year) | In-migration (relative attractivity as destination) |
| --- | --- | --- |
| Chitipa | 1.0% | 1 |
| Karonga | 1.0% | 1 |
| Likoma | 1.5% | 1 |
| Mzimba | 1.0% | 1 |
| Nkhata Bay | 1.2% | 1 |
| Rumphi | 1.2% | 1 |
| Dedza | 2.0% | 1 |
| Dowa | 2.0% | 1 |
| Kasungu | 1.2% | 1 |
| Lilongwe | 1.0% | 1 |
| Mchinji | 1.2% | 1 |
| Nkhotakota | 1.5% | 1 |
| Ntcheu | 2.0% | 1 |
| Ntchisi | 1.5% | 1 |
| Salima | 1.0% | 1 |
| Balaka | 1.5% | 1 |
| Blantyre | 1.5% | 1 |
| Chikwawa | 1.5% | 1 |
| Chiradzulu | 1.5% | 1 |
| Machinga | 1.0% | 1 |
| Mangochi | 1.0% | 1 |
| Mulanje | 1.5% | 1 |
| Mwanza | 1.5% | 1 |
| Nsanje | 1.5% | 1 |
| Thyolo | 1.5% | 1 |
| Phalombe | 1.2% | 1 |
| Zomba | 2.0% | 1 |
| Neno | 1.0% | 2 |

Unprotected sex acts per year

|  | Model I | Model II | Model III | Model IV | Model V | Model I | Model II | Model III | Model IV | Model V |
| --- | --- | --- | --- | --- | --- | --- | --- | --- | --- | --- |
| 1975 | 20.1894 | 20.6144 | 20.9332 | 20.6144 | 21.252 | 68.2594 | 69.6964 | 70.7742 | 69.6964 | 71.852 |
| 1976 | 20.1894 | 20.6144 | 20.9332 | 20.6144 | 21.252 | 68.2594 | 69.6964 | 70.7742 | 69.6964 | 71.852 |
| 1977 | 20.1894 | 20.6144 | 20.9332 | 20.6144 | 21.252 | 68.2594 | 69.6964 | 70.7742 | 69.6964 | 71.852 |
| 1978 | 20.1894 | 20.6144 | 20.9332 | 20.6144 | 21.252 | 68.2594 | 69.6964 | 70.7742 | 69.6964 | 71.852 |
| 1979 | 20.1894 | 20.6144 | 20.9332 | 20.6144 | 21.252 | 68.2594 | 69.6964 | 70.7742 | 69.6964 | 71.852 |
| 1980 | 20.1894 | 20.6144 | 20.9332 | 20.6144 | 21.252 | 68.2594 | 69.6964 | 70.7742 | 69.6964 | 71.852 |
| 1981 | 20.1894 | 20.6144 | 20.9332 | 20.6144 | 21.252 | 68.2594 | 69.6964 | 70.7742 | 69.6964 | 71.852 |
| 1982 | 20.1894 | 20.6144 | 20.9332 | 20.6144 | 21.252 | 68.2594 | 69.6964 | 70.7742 | 69.6964 | 71.852 |
| 1983 | 20.1894 | 20.6144 | 20.9332 | 20.6144 | 21.252 | 68.2594 | 69.6964 | 70.7742 | 69.6964 | 71.852 |
| 1984 | 20.1894 | 20.6144 | 20.9332 | 20.6144 | 21.252 | 68.2594 | 69.6964 | 70.7742 | 69.6964 | 71.852 |
| 1985 | 20.1894 | 20.6144 | 20.9332 | 20.6144 | 21.252 | 68.2594 | 69.6964 | 70.7742 | 69.6964 | 71.852 |
| 1986 | 20.1894 | 20.6144 | 20.9332 | 20.6144 | 21.252 | 68.2594 | 69.6964 | 70.7742 | 69.6964 | 71.852 |
| 1987 | 20.1894 | 20.6144 | 20.9332 | 20.6144 | 21.252 | 68.2594 | 69.6964 | 70.7742 | 69.6964 | 71.852 |
| 1988 | 20.1894 | 20.6144 | 20.9332 | 20.6144 | 21.252 | 68.2594 | 69.6964 | 70.7742 | 69.6964 | 71.852 |
| 1989 | 20.1894 | 20.6144 | 20.9332 | 20.6144 | 21.252 | 68.2594 | 69.6964 | 70.7742 | 69.6964 | 71.852 |
| 1990 | 20.1894 | 17.1108 | 17.3754 | 17.1108 | 21.252 | 68.2594 | 57.8508 | 58.7454 | 57.8508 | 71.852 |
| 1991 | 20.1894 | 17.1108 | 17.3754 | 17.1108 | 15.939 | 68.2594 | 57.8508 | 58.7454 | 57.8508 | 53.889 |
| 1992 | 15.96 | 17.1108 | 17.3754 | 17.1108 | 15.939 | 53.96 | 57.8508 | 58.7454 | 57.8508 | 53.889 |
| 1993 | 15.96 | 17.1108 | 17.3754 | 17.1108 | 15.939 | 53.96 | 57.8508 | 58.7454 | 57.8508 | 53.889 |
| 1994 | 15.96 | 17.1108 | 17.3754 | 17.1108 | 15.939 | 53.96 | 57.8508 | 58.7454 | 57.8508 | 53.889 |
| 1995 | 15.96 | 15.4812 | 15.7206 | 15.4812 | 15.939 | 53.96 | 52.3412 | 53.1506 | 52.3412 | 53.889 |
| 1996 | 10.0947 | 15.4812 | 15.7206 | 15.4812 | 15.939 | 34.1297 | 52.3412 | 53.1506 | 52.3412 | 53.889 |
| 1997 | 10.0947 | 15.4812 | 15.7206 | 15.4812 | 15.939 | 34.1297 | 52.3412 | 53.1506 | 52.3412 | 53.889 |
| 1998 | 10.0947 | 15.4812 | 15.7206 | 15.4812 | 15.939 | 34.1297 | 52.3412 | 53.1506 | 52.3412 | 53.889 |
| 1999 | 10.0947 | 15.4812 | 15.7206 | 15.4812 | 15.939 | 34.1297 | 52.3412 | 53.1506 | 52.3412 | 53.889 |
| 2000 | 10.0947 | 13.4442 | 12.411 | 13.4442 | 15.939 | 34.1297 | 45.4542 | 41.961 | 45.4542 | 53.889 |
| 2001 | 10.0947 | 13.4442 | 12.411 | 13.4442 | 15.939 | 34.1297 | 45.4542 | 41.961 | 45.4542 | 53.889 |
| 2002 | 10.0947 | 13.4442 | 12.411 | 13.4442 | 10.626 | 34.1297 | 45.4542 | 41.961 | 45.4542 | 35.926 |
| 2003 | 11.97 | 13.4442 | 12.411 | 13.4442 | 10.626 | 40.47 | 45.4542 | 41.961 | 45.4542 | 35.926 |
| 2004 | 11.97 | 12.222 | 12.411 | 12.222 | 10.626 | 40.47 | 41.322 | 41.961 | 41.322 | 35.926 |
| 2005 | 11.97 | 12.222 | 12.411 | 12.222 | 10.626 | 40.47 | 41.322 | 41.961 | 41.322 | 35.926 |
| 2006 | 11.97 | 12.222 | 12.411 | 12.222 | 10.626 | 40.47 | 41.322 | 41.961 | 41.322 | 35.926 |
| 2007 | 11.97 | 12.222 | 12.411 | 12.222 | 10.626 | 40.47 | 41.322 | 41.961 | 41.322 | 35.926 |
| 2008 | 11.97 | 12.222 | 12.411 | 12.222 | 10.626 | 40.47 | 41.322 | 41.961 | 41.322 | 35.926 |
| 2009 | 11.97 | 12.222 | 12.411 | 12.222 | 10.626 | 40.47 | 41.322 | 41.961 | 41.322 | 35.926 |
| 2010 | 11.97 | 12.222 | 12.411 | 12.222 | 10.626 | 40.47 | 41.322 | 41.961 | 41.322 | 35.926 |
| 2011 | 11.97 | 12.222 | 12.411 | 12.222 | 10.626 | 40.47 | 41.322 | 41.961 | 41.322 | 35.926 |
| 2012 | 11.97 | 12.222 | 12.411 | 12.222 | 10.626 | 40.47 | 41.322 | 41.961 | 41.322 | 35.926 |
| 2013 | 11.97 | 12.222 | 12.411 | 12.222 | 10.626 | 40.47 | 41.322 | 41.961 | 41.322 | 35.926 |
| 2014 | 11.97 | 12.222 | 12.411 | 12.222 | 10.626 | 40.47 | 41.322 | 41.961 | 41.322 | 35.926 |
| 2015 | 11.97 | 12.222 | 12.411 | 12.222 | 10.626 | 40.47 | 41.322 | 41.961 | 41.322 | 35.926 |
| 2016 | 11.97 | 12.222 | 12.411 | 12.222 | 10.626 | 40.47 | 41.322 | 41.961 | 41.322 | 35.926 |
| 2017 | 11.97 | 12.222 | 12.411 | 12.222 | 10.626 | 40.47 | 41.322 | 41.961 | 41.322 | 35.926 |
| 2018 | 11.97 | 12.222 | 12.411 | 12.222 | 10.626 | 40.47 | 41.322 | 41.961 | 41.322 | 35.926 |
| 2019 | 11.97 | 12.222 | 12.411 | 12.222 | 10.626 | 40.47 | 41.322 | 41.961 | 41.322 | 35.926 |
| 2020 | 11.97 | 12.222 | 12.411 | 12.222 | 10.626 | 40.47 | 41.322 | 41.961 | 41.322 | 35.926 |

**Figure S1. Average annual unprotected sex acts with casual partners in the models I to V.** See Table 1 of the main text for a definition of the models. Solid lines present the low risk group and dashed lines the high risk group.
